# Music presentation modulates metabolic and physiologic condition of patients in the ICU

**DOI:** 10.64898/2025.12.31.25343291

**Authors:** Jagmeet S. Kanwal, Josephine Millard, Andrew Schulman, Abigail Perelman, Preeti Kota, Anna J. Patel, Julia Langley

**Affiliations:** Department of Neurology, Georgetown University Medical Center, Washington, DC. 20057; Georgetown Lombardi Arts and Humanities Program, Georgetown University Medical Center, Washington, DC. 20057; Medical Musician Initiative, New York, NY, 10025; Department of Psychology, Georgetown University, Washington, DC. 20057; Department of Biology, Georgetown University, Washington, DC. 20057

**Author notes:** Correspondence author and address: Department of Neurology Georgetown University Medical Center, 3900 Reservoir Road, NW, Washington DC, 20057-1460, USA.

**Keywords:** music, heart rate, stress, ICU, liver transplant, autonomic activity, skin conductance, cortisol

## Abstract

The embodied brain is highly dynamic, changing with every thought, sensory input and motor activity. It keeps us coherent and healthy via its connections to every organ within the body, particularly the heart, which in turn supplies nutrients and oxygen to all bodily organs and the brain. Listening to music can instantly alter brain-body dynamics. Yet, the acoustic, neural, and physiologic parameters and processes that facilitate these effects are not well understood. Here, we tested the hypothesis that a custom music composition can promote healing in patients recovering from liver transplant surgery within an intensive care unit (ICU). The music presented consisted of custom,15-minute music sets curated and recorded by an experienced medical musician. We obtained cortisol samples from saliva samples ∼15 minutes before and after music presentation and captured autonomic activity by recording electrocardiography for 5 minutes before, during, and 5 minutes after music presentation in normal subjects and patients. Discriminant analysis showed a significant decrease in cortisol production (n = 17) after music presentation. Detailed analysis in a single patient showed significant changes in multiple cardiac parameters, including heart-rate variability (HRV). Multidimensional scaling of twenty-five parameters related to HRV in a patient mapped all five instances of the music presentation condition outside of the mixed cluster of baseline conditions before and after music presentation. Our results show that listening to music promotes homeostasis in ICU patients by transiently shifting physiological parameters towards a state of recovery that may stabilize over time.

## 1. Introduction

As the depth of our understanding of the interaction between the mind/brain and the body increases, we can develop new interventions for alleviating mental and physiological dysfunctions. For example, deep brain stimulation and other electroceutical treatment methodologies are already beginning to provide alternate and often more effective means of treatments (Trost et al., 2018; van Vugt et al., 2013). These unconventional and complimentary approaches have the distinct advantage of minimizing undesirable side-effects that are a universal feature of pharmaceutical approaches. Brain stimulation, however, can be realized in more natural, albeit subtle, ways through brain pathways for sensory perception. Thus, intense olfactory stimuli are known to reduce epileptic seizure susceptibility and even block its onset (Delfino-Pereira et al., 2018; Ebert and Löscher, 2000). Similarly, there are widely circulated anecdotal and experimentally tested instances of music enabling a long lasting positive change in the condition of critically ill patients critically ill patients (Pant et al., 2022; Sacks, 2008; Schulman, 2016). These together with an increasing number of scientific studies on the therapeutic and physiological effects of music have elevated its role within the context of healing and medicine (Raglio and Oasi, 2015). They include the use of music as a tool for alleviating or minimizing pain during and after medical procedures, including childbirth (Bernatzky et al., 2011; Browning, 2000; Hartling et al., 2013; Yaman Aktaş and Karabulut, 2019). This analgesic effect is believed to be mediated through the release of endorphins and other neurochemical changes in the brain, which can diminish the need for pharmacological pain management and its associated side effects (Mitchell et al., 2008).

Studies on the therapeutic effect of music on various physiological and psychological parameters also include its ability to regulate heart rate. Listening to calming music even in normal subjects can lead to a significant reduction in heart rate, likely due to its influence on the autonomic nervous system (Elliott et al., 2011). This effect is particularly beneficial for patients recovering from surgery or dealing with chronic health conditions where heart rate variability is a key indicator of overall cardiovascular health (Escher and Evéquoz, 1999; Koelsch and Jäncke, 2015; Trappe, 2010). In terms of probing the brain, music can also restore memories, albeit transiently, in patients suffering from Alzheimer’s as well as allow shell-shocked and Parkinson’s patients to move to a piece of music are well documented (Boso et al., 2006; Clark and Warren, 2015; Izbicki et al., 2020).

Music also plays a critical role in stress management, as evidenced by its impact on cortisol levels, a primary biomarker of stress. Music’s ability to lower cortisol and other stress-related hormones can help create a more stable internal environment, promoting healing and reducing the risk of complications related to prolonged stress. Moreover, music presentation has been shown to alleviate anxiety, providing a non-invasive means of improving mental well-being. The use of music to reduce anxiety can be especially beneficial in the context of post-operative care, where patients often experience heightened levels of anxiety due to pain, uncertainty, and the challenging nature of recovery (Chanda and Levitin, 2013). These effects are likely mediated by the complex interplay between the brain’s emotional centers, such as the amygdala and hypothalamus, and the autonomic nervous system, which regulates immune function. By inducing a state of relaxation and well-being, music may create a favorable environment for immune system optimization. These multifaceted effects of music compelled us to examine music induced enhancement of recovery from liver transplant surgery in patients admitted to an Intensive Care Unit (ICU).

Liver transplant surgery is a complex and life-saving procedure involving the replacement of a diseased or failing liver with a healthy donor organ. This surgery is typically reserved for patients with end-stage liver disease or acute liver failure, where other treatments have failed. In the United States alone, there are over 8,000 liver transplants performed annually, reflecting the significant burden of liver disease in the population(Demir et al., 2021). The speed and quality of recovery from surgery, particularly during the initial phase when the patient is recovering in the ICU, has a critical impact on the overall success of liver transplant surgery and prognosis over the long-term for patients. During this period, the patients need to be closely monitored for signs of organ rejection, infection, and other complications. This period often involves the administration of immunosuppressive drugs, careful management of fluid and electrolyte balance, and the gradual reintroduction of nutrition. Recovery from the effects of anesthesia typically occurs within a few hours after the surgery, but the timeline can vary depending on the individual’s overall health, the type of anesthesia used, and the complexity of the procedure. Most patients regain consciousness within an hour or two after surgery, but they may continue to experience grogginess, confusion, or disorientation for several hours as the anesthesia wears off completely. Older adults and those suffering from a complication, can also suffer from postoperative delirium that can last from a few days to several weeks. Typically, however, patients can be discharged from the ICU in two to three days.

Given the physical and psychological challenges inherent in post-transplant recovery, particularly in the ICU setting, there is growing interest in non-pharmacological interventions to enhance patient outcomes. If music can either induce or enhance recovery, then it has the potential to conserve resources by decreasing time to discharge, probability of complications, and return visits, lowing the financial burden of post-transplant recovery for both hospitals and patients. In this context and considering the work of others detailed above, we propose that listening to music can play a role in reducing anxiety, alleviating pain, and promoting overall well-being during the arduous recovery process. To test our hypothesis, we needed to detect an observable effect of music presentation. Since each patient’s situation is unique, it was not possible to have viable controls or double-blind administration of a treatment, i.e., of music. In a large-scale study, an evaluation of the specific effects of music presentation can be controlled with presentation of other types of sounds and even different types of music. In this first of its kind study, however, we chose to simply test if the presentation of music had any impact at all on the vital physiological parameters, such as heart-rate variability (HRV) and stress levels. By obtaining data prior to, during and post-music presentation, each patient acted as their own control as we tested for measurable changes reflecting body and brain dynamics. These changes were taken to be signs of the effect of music and the potential for improved recovery. We also made similar measurements, using the same music, in a population of healthy subjects to test whether the observed effects are grounded in normal physiology and not unique to the ICU condition. Together, our data support the presence of universal effects of listening to music that could prove beneficial to patients in the ICU by promoting recovery. At this stage, we cannot claim that they result in long-lasting effects or enhance recovery. To do so will require a large-scale, longitudinal study/clinical trial and a larger investment of time and funds than what was presently available.

A relatively unique aspect of our study was the presentation of a set of musical pieces, created and recorded especially for this study by a medical musician (https://andrewschulmanmusic.com/about). These musical pieces were arranged based on a deep understanding of the healing process from many years of experience in the practice of presentation of music to patients. The pieces of music, termed here as “penicillin pieces” were designed to both calm and arouse a patient from a state of delusion and stress that typically results from transplant surgery. Each piece was created for presentation at a different time of the day.

## 2. Methods

### 2.1. Study setting, eligibility and recruitment

The research took place in the liver transplant ICU of Medstar-Georgetown University Hospital (MGUH), where continuous monitoring and measurement of physiological parameters in patients is routine. The study team together with the nursing staff monitored ICU patients for clinical indicators of any discomfort during and immediately after the patient listened to the music. Our interdisciplinary team included researchers, clinicians and arts practitioners with a breadth and depth of expertise and has a long history of investigating the neurologic effects of emotive sounds, including music and its practice with other arts to alleviate suffering in the clinical environment. For music presentation in normal human subjects, we recruited mostly undergraduate students at the university campus. The constitution of the subject was Caucasian/White (60%) Asian (30%) and Other/No Answer (10%) per the voluntary questionnaire provided. Mean age of 6 males and 10 females included in the study was 27.2 ± 15.138.

We recruited patients between the ages of 18 to 75 from the MGUH Transplant Institute admitted from a pool of outpatient, post-liver transplant individuals. Patients who needed intubation or were heavily sedated (RASS < -2) or those expressing extreme agitation (RASS > 2) were excluded from the study. We used RASS, the Richmond Agitation-Sedation Scale, to quantify the level of sedation (-5 is unarousable, +4 is extreme agitation, 0 is alert and calm).

Patients who satisfied the inclusion/exclusion criteria and expressed an interest in participating in the study were contacted either prior to or during the admission process to inform them about the opportunity to receive therapeutic music during their stay in the recovery room. Those contacted by phone filled out a consent form and a questionnaire to collect pre-therapy data prior to hospital admission. They were also provided a link to samples of the music. Participants’ musicality, motivation for music use, and anxiety levels were also assessed to evaluate if and how musicality and mental state correlate with individual physiologic changes. Musicality was scored using the standardized music background questionnaire (MUSEBAQ). Music presentation schedule was established in consultation with the nursing station staff. Admitting personnel, attending physicians, as well as floor nurses, as appropriate, were informed and educated about the project prior to its initiation. All methods were approved by the Institutional Review Board (IRB).

### 2.2. Experimental design

#### 2.2.1. Music preparation and presentation

Baseline data on the time course of normal patterns of recovery in patients in a typical ICU care setting at MGUH were used to determine appropriate times for music presentation, i.e., when physiologic parameters are relatively stable. Music was played morning, mid-day, and in the evening. The music pieces were organized, played on an eight-string solo guitar by a certified therapeutic musician (Andrew Schulman) and recorded in Ted Spencer Recording Studio, a professional studio in New York City, NY) (an eight-string guitar has two extra strings in the bass, and the extra depth of sound is very conducive to healing sounds). The selection of the music pieces was driven to induce a state of soothing and healing in the subjects. The basic parameters for choosing therapeutic music, designated by Schulman as “Penicillin pieces”, consisted of tempos that were moderate and steady, as well as melodic/lyrical, utilizing variety in the sequencing of musical pieces. Moderate tempos were deemed ideal for matching the normal heart rate range of 60-100 bpm. Melodic/lyrical music were designed to stimulate the patient’s imagination during playback, focusing on a blossoming effect of music. Variety in repertoire, was achieved by matching selections that included both classical and popular music. This was considered vital for keeping the patient engaged when listening to music. In the three sets (see list below) played to patients. Music was presented during morning, mid-day, and in the evening. The music consisted of seven classical pieces, seven popular pieces, and one original work

##### Morning Set

Prelude in D, BWV 1007…J.S. Bach

Here Comes the Sun…George Harrison

The Girl with the Flaxen Hair…Claude Debussy

The Entertainer, from The Sting…Scott Joplin

Little Theme in D…Peter Williams

##### Noon Set

Jesu, Joy of Man’s Desiring, BWV 147…Bach

Adagio in C…Mozart

Someone to Watch Over Me…Gershwin

Here, There and Everywhere…Lennon/McCartney

Cavatina, from The Deer Hunter…Stanley Myers

##### Evening Set

Prelude in D minor, BWV 999…Bach

La Vie en Rose…Piaf/Louiguy

Julia…Lennon/McCartney

Elegy…Andrew Schulman

Somewhere Over the Rainbow, from The Wizard of Oz…Arlen/Harburg

In the three sets of music played, the mood was established by the first piece, in each case a short work by Bach. For example, the Morning Set begins with a *Prelude* intended to slowly lead to an awakening, and is followed by a well-known popular tune, *Here Comes the Sun*, to reinforce that mood. In the Noon Set the well-known *Jesu, Joy of Man’s Desiring* had a flowing pulse in the upper voice and a steady bass underpinning the melody – music that is fully engaging. In the Evening set the *Prelude* was ruminative music giving a sense of the end of the day; to patients, the mood was established by the first piece in each case.

#### 2.2.2. Procedural details

During registration at the Surgical or Medical ICU desk patients completed a trait and state anxiety inventory (STAI) that is a commonly used measure of adult trait and state anxiety (38). Once the consent form was received, the research team, attending physician and floor staff, were notified that the patient has consented to receiving the music intervention. Within 24 hours of the patient reaching the surgical ward of the ICU, the sound system was set up in the patient’s room by wheeling in the speaker and placing it at the feet of the patient’s bed so that it faced the patient. We also used the Music Use and Background Questionnaire (MUSEBAQ), a self-report instrument designed to measure the level of music engagement and background for each subject/patient. These data allowed us to test if this measure correlated with any of the changes in anxiety or physiological parameters after music presentation as well as obtain an estimate of the heterogeneity of the subject population available for the study.

### 2.3. Data collection and analysis

Speakers were tested and their frequency-response curve obtained within a soundproof room for calibrating sound output across a wide (20 Hz to 20 kHz frequency-range). Background noise was recorded with Audacity software. The best sound pressure level was determined to be ∼ 70 dB SPL. Music was played back through a high-quality speaker (Beosound A9, Bang & Olufsen, Inc.), which reproduced the lower frequencies in a piece of music particularly well. Ambient sound level was measured and recorded, and the quality of sound output was checked and calibrated. The patient was asked if they could hear the sound, and whether the sound level was within a comfortable range. Ambient noise levels were kept to a minimum (< 40 dB).

The AIM galvanic skin response device was attached to the finger to measure levels of anxiety and arousal. A saliva sample was obtained at the beginning of music presentation and again within 30 min after the end of a 15-minute music presentation. This procedure was repeated for mid-day and evening music presentations. Between presentations, the sound system and experiment equipment were transported out of the room of the patient-participant. A questionnaire was filled out with the assistance of a nurse just prior to patient discharge.

The schematic in figure 1 shows the timeline for data collection from all subjects during this study, including EEG data in a subset of the subjects. Cortisol was sampled from saliva collected ∼15 minutes before and after music presentation. Cortisol swabbing was performed as prescribed by Saltimetrics, Inc. where doublet sample assays were also completed. Data from AIM were streamed to a dedicated laptop computer and archived. This system also captured autonomic activity by recording electrocardiographic, respiration, photoplethysmography, temperature, and electrodermal activity for 5 minutes before, during, and 5 minutes after music presentation. The electrodermal activity tracked the galvanic skin response (GSR) to measure levels of anxiety and arousal and dynamic air pressure changes. We used MNE & Neurokit2 Python packages to analyze physiological data, such as heart rate from an electrocardiogram (EKG), breathing rate, and skin conductance changes.

**Figure 1.**
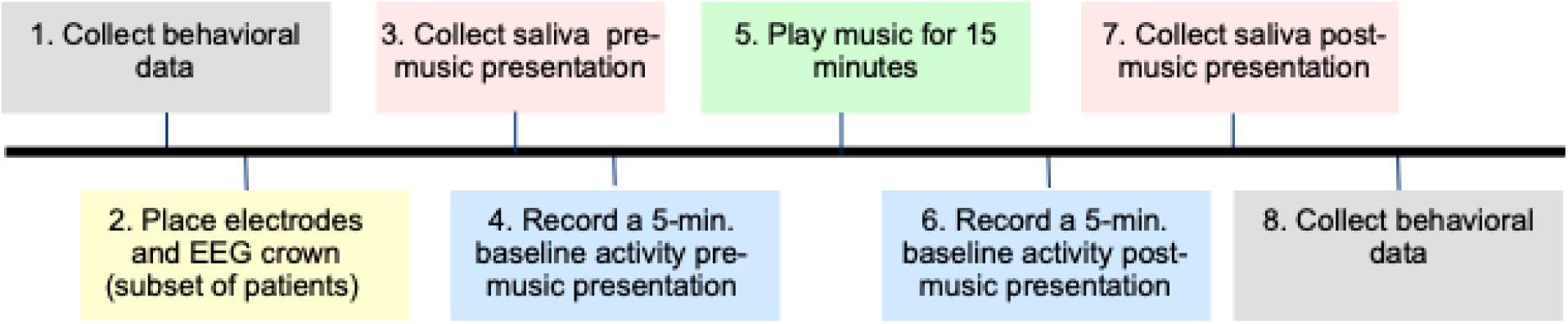
Timeline for data acquisition from patients and healthy subjects. For ICU patients, music was presented 3 times (morning, early afternoon and evening.

**For** data analysis, music played to the patient was re-recorded in synchrony with other data streams and then correlated with the patient’s physiological parameters, including dynamic correlation with the GSR and SPP. Simultaneous recording of electrocardiographic, respiration, photoplethysmographic, temperature, and electrodermal activity (AIM3, Brain Vision) were streamed to a laptop computer, and archived. This system captured autonomic activity by recording. This system captured autonomic activity by recording the activity for 5 minutes before, during, and 5 minutes after music presentation. The metabolic profile included measurement of cortisol in saliva to provide a window into the instantaneous effects of music on vital physiologic parameters. From the EEG data, we tested for a negative correlation between alpha waves and cortisol levels

### 2.4. Data management and statistical methods

All patient information was de-identified and protected per HIPAA regulations. Aggregate data, analyses, and plots were made using statistical and graphical software (JMP-pro, ver. 8.1, SAS Inc. and Canvas, Deneba, Inc.). All data was archived using “Georgetown Box” that is a HIPPA/PH approved cloud-based service.

We conducted statistical analysis after organizing subject-coded data and performing outlier analysis (Quantile method) and removing any outliers through two stages of filtering. We performed classical (frequentist) statistical analyses to test for significance (P<0.05) of changes in the cortisol profile after listening to music. Simple linear regression was used to test for effects of music presentation on single variables such as music preference and individual HRV parameters. We used nonparametreic methods to systematically analyze individual variation within each of the recorded parameters, such as cortisol and heart-rate variability (HRV). Physiological normality represents one homogeneous state of subjects from which multiple physiological parameters in patients drift away due to their illness, leading to an increase in between-subject variance. This drift away from normality creates a more heterogenous pool of physiological data. Therefore, if a treatment such as music presentation triggers a return-of-function effect, then one would expect the overall status to move towards a more homogenous state, representing the normal condition. One way to gauge improvement is to test whether music presentation reduces the extent of multiparametric variation between subjects. This is more likely to correspond to the normal condition. To test this effect, we performed multiparametric discriminant analysis (DA) using the regularized compromise method that yielded a good fit (0% misclassification an entropy R^2^value of >0.096), during and after presentation of music. We performed a similar analysis across multiple trials of music presentation to test the cumulative effect of music presentation. We could not conclude or evaluate the beneficial effect of music without longitudinal tracking of patients in a randomized clinical trial-like manner, given the small cohort of subjects. We could test, however, whether there was any uniformity in the effect of music on HRV parameters. All music presentation trials and measured parameters were pooled for this analysis.

Since each subject is unique in their physical condition, the level of stress post-surgery as well as genetic and mental traits, we picked a single subject with the maximum number of presentations of music and examined how the presentation of music changed the HRV parameters as a case study. We examined the placement of multiple HRV parameters in a reduced (to 2 dimensions for visualization purposes) multidimensional space using multidimensional scaling (MDS) procedures (JMPpro ver. 8.1). This approach can yield insights into the mechanism of action of a drug or a treatment for rare or unique conditions. We adopted this approach in one subject for a to illustrate a consistent effect of music presentation and gaining insights into how music might contribute to recovery and return of function in liver transplant subjects. We then examined the data in additional patients receiving fewer (3 to 6) trials of music presentation as an intervention).

## 3. Results

We focus here mainly on the effect of music presented to liver transplant patients in the ICU. As a comparison for an effect of music in a population of normal subjects, we also include results of those experiments here. This also provided a test for optimal functioning of the recording apparatus and trouble-shooting any hardware/software issues as the ICU was not was not an optimal environment for doing so. For this reason, recordings in normal subjects preceded and alternated, when possible, with presentation of music in the ICU.

### 3.1. Role of musicality

Analysis of data obtained for 11 parameters from the MUSEBAQ questionnaire indicated a greater heterogeneity among the patient population compared to the normal subject population. A Random Forest clustering analysis revealed a best fit (lowest BIC criterion) for both normal human subjects (n = 29) and patients (n = 10) revealed two clusters. The two clusters were not separated by the sex of the subjects in either case. Of the various parameters, “Emotional sensitivity to music”, “Music memory and imagery” and “Personal commitment to music” criteria were the most highly correlated (*r* = 0.53 to 0.76) with STAI state anxiety scores.

Data on musical capacity, and music use reported through the MUSEBAQ questionnaire revealed that individual changes in cortisol production over the course of music presentation were significantly correlated (P = 0.0218; n = 7) with self-reported use of music as a tool for cognitive regulation (Fig. 4). There was no difference among males and females for normal subjects before and after music presentation (P = 0.95 and 0.79; females = 16 and males = 10).

### 3.2. Effect on cortisol levels and anxiety

Across both populations, of normal, healthy, subjects and patients, cortisol levels in males and females were not significantly different. In normal healthy subjects, baseline cortisol levels on average were 0.250 µg/dL in both females and males, whereas in patients, the mean values of cortisol levels were 0.427 µg/dL in females and 0.398 µg/dL in males. This indicated that ICU patients had nearly double the level of cortisol level compared to that of normal healthy subjects (Fig. S1). This is understandable given that ICU patients had undergone liver transplant surgery. A breakdown of changes in salivary cortisol levels by subject population and sex provided mixed effects of music presentation. This result is shown as box plots for all healthy subjects in figure 2A and compared to those for males alone (Fig. 2B). Across all subjects of both sexes, music presentation resulted in a slight decrease in cortisol levels (0.1792 ± 0.096 to 0.166 ± 0.103; P = 0.169 µg/dL for Kruskal-Wallis rank sum test assuming unequal variances; n = 56 samples in 28 subjects). The decrease, however, was significant for males (P < 0.05 for Student’s paired t-test; data were normally distributed with no significant differences between subjects). On average, cortisol levels in males decreased by ∼0.05 µg/dL within within a 20 to 30 minute interval.

**Figure 2:**
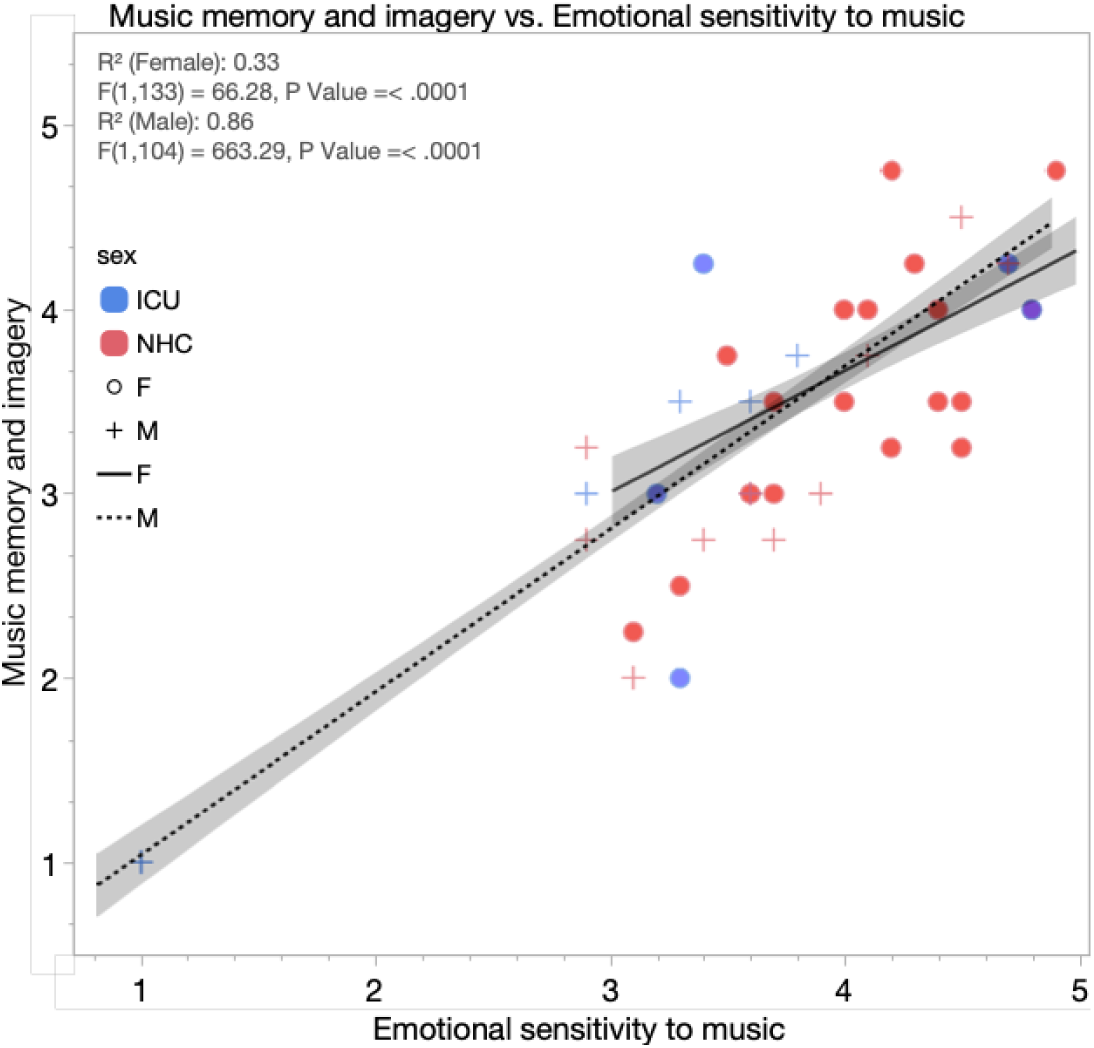
Line plot to show correlatin between “Music memory and imagery” and Emotional sensitivity to music”variables in the MUSEBAQboth of which were highly correlated with STAI-state anxiety scores in both normal subjects and ICU patients. The plot shows a sex difference in that males showed a much higher correlation than females for these two parametrs. This may explain the larger decrease in cortisol levels observed in males compared to females (see Fig 4).

In the ICU patients, presentation of music resulted in a slight but significant increase (from a mean ± stand. dev. of 0.414 ± 0.292 to 0.494 ± 0.329 µg/dL; P < 0.001 for Kruskal-Wallis rank sum test assuming unequal variances, n = 67 samples in 15 subjects) in the level of salivary cortisol (Fig. 3A). This increase was relatively consistent across subjects, as well as in both sexes. The increase in males was < 0.06 µg/dL. Model fit showed significant (P < 0.05) differences between subjects. Closer analysis revealed a relatively high baseline level of cortisol levels, understandably so, because of the stress associated with liver transplant surgery. Very low levels (< 0.30 µg/dL) obtained in some patients could have resulted from inadequate saliva samples or from lingering effects of anesthesia. For middle to high ranges of cortisol levels (> 0.30 µg/dL) the level of cortisol levels in both sexes declined dramatically (P < 0.0001, Kruskal-Wallis rank sum test; from a mean of 0.87 to 0.48 µg/dL in males) after music presentation (Fig. 3B). The decrease in females was a little lower (0.32 µg/dL) but still significant.

**Figure 3:**
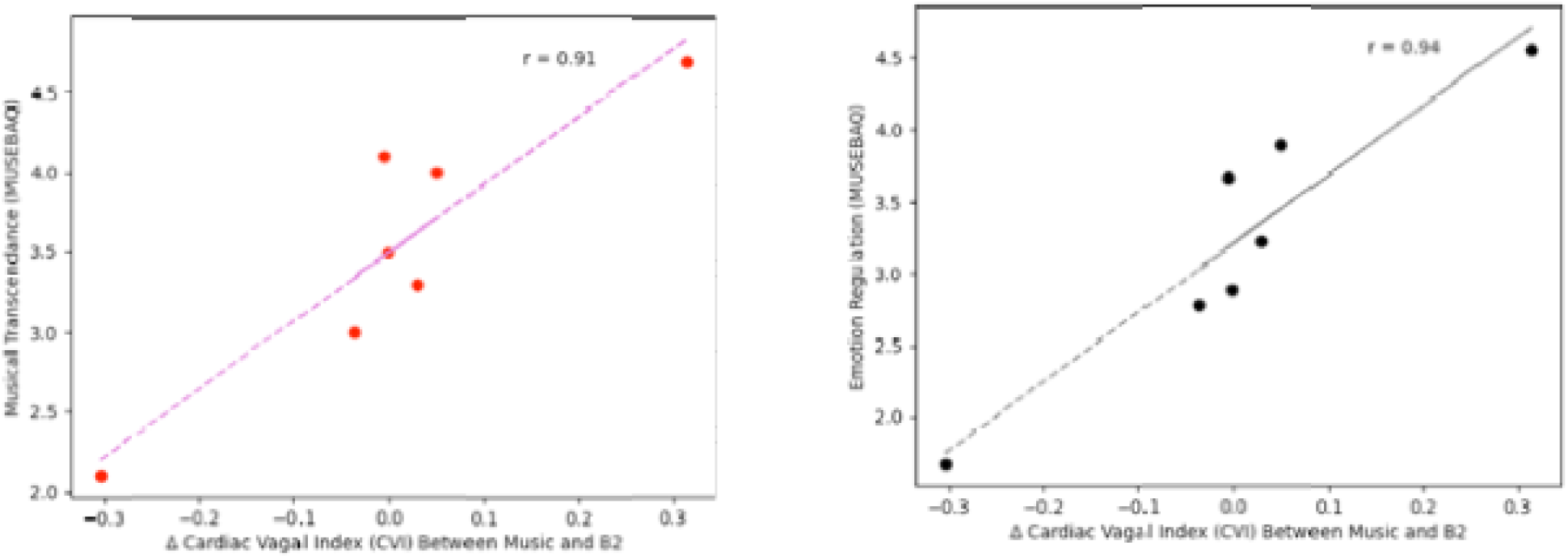
Degree of motivation for music use measured via a musicality questionnaire in patients before music presentation. Music presentation was significantly correlated with the change in parasympathetic outflow measured as cardiac vagal index via EKG (n = 7, Emotion Regulation: p = 0.0019, musical transcendence: P = 0.0042). Cardiac Vagal Index is assumed to be an indicator of cardiac parasympathetic function and expected to be unaffected by sympathetic activity.

Questionnaire data collected on anxiety provided an average STAI trait score of 38.52±10.31 (n = 7) and an STAI state score of 44.84±11.85 in ICU patients. In normal subjects, the STAI trait score was 39.5 ± 11.378 (n = 16) and the state score was 33.41. Clearly, despite having similar trait anxiety, ICU patients were in a higher state of anxiety compared to normal subjects, which can be reasonably expected. The initial STAI State score decreased by ∼5 points post-music presentation in ICU patients and by ∼ 9 points in post-music presentation in normal subjects. There was no clear relationship, however, between changes in state anxiety scores and cortisol levels for normal subjects where data were available (P = 0.822).

### 3.3. Heart rate parameters

To study the effect of music presentation on the heart, we focused on multiple HRV parameters extracted using Python kit. This provided a way to test the sympathetic/parasympathetic balance of activity as expressed in the timing of successive heartbeats. Therefore, for a more general representation of within-subject data to the overall effects of music presentation on HRV, we examined the separation across subjects in the multifactorial HRV space using DA (Fig. 6A). These data showed that there is a large degree of separation between subjects (n = 9) in the HRV space for music presentation and the before and after intervals as the discriminant variable. Canonical details revealed that the first eigenvalue captured 93 to 96% of the variation and showed a good fit (P < 0.0001; F = -13.32, 440, -219 for the first DA component). The presentation of music reduced the canonical distance between subjects on the first canonical axis such that it was more difficult to discriminate their physiological condition (HRV status) during music presentation vs. the baseline condition music. Post-music presentation, there was some separation in the discriminant space, but the subjects mapped relatively closely to each other compared to the pre-music presentation, suggesting that music presentation tended to modify HRV parameters across subjects towards a common status during music presentations.

**Figure 4:**
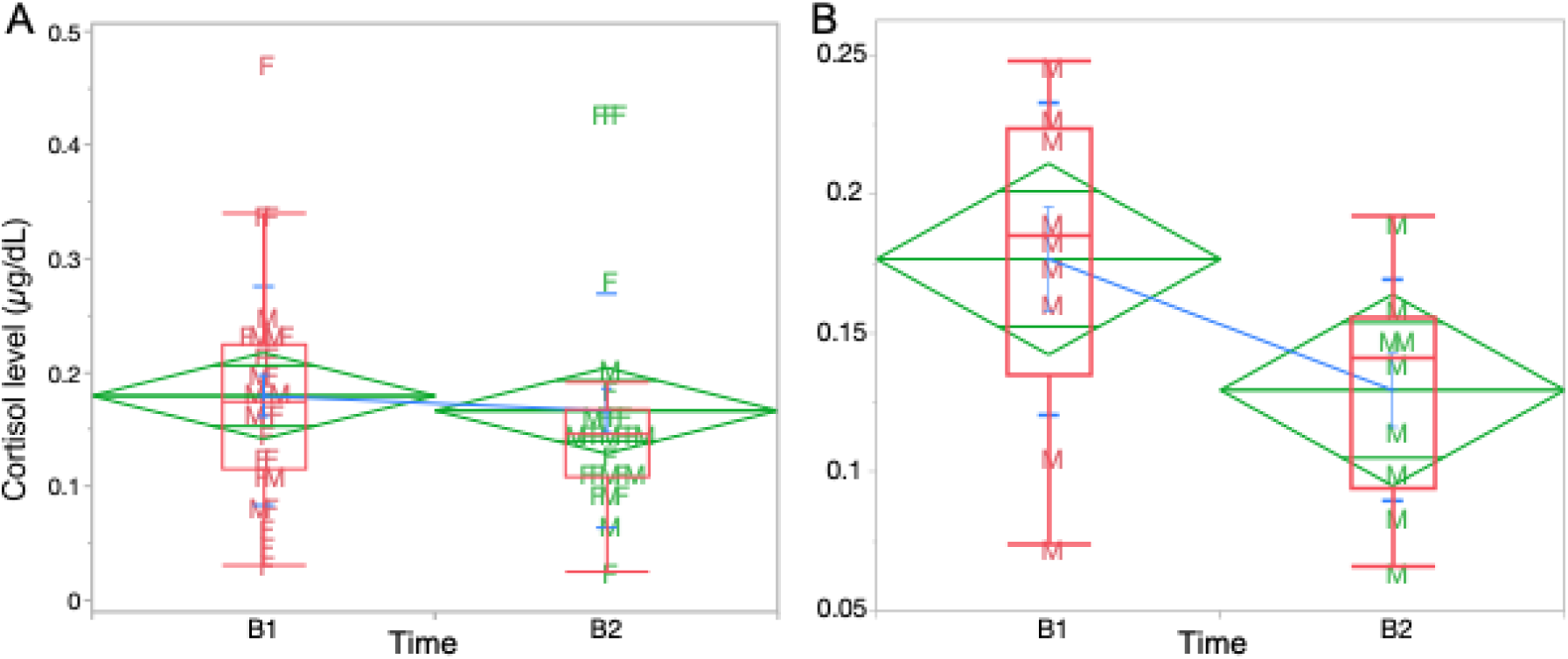
Box plots showing distribution and comparison of changes in salivary cortisol levels before (B1) and after (B2) music presentation in (A) all healthy subjects and (B) in males (M). Red horizontal lines indicate median leavels and green diamonds represent 95% confidence intervals from the pooled stanbdard deviation. Bue line connects group means (long horizontal green lines). Short horizontal lines indicated standard deviation.

**Figure 5:**
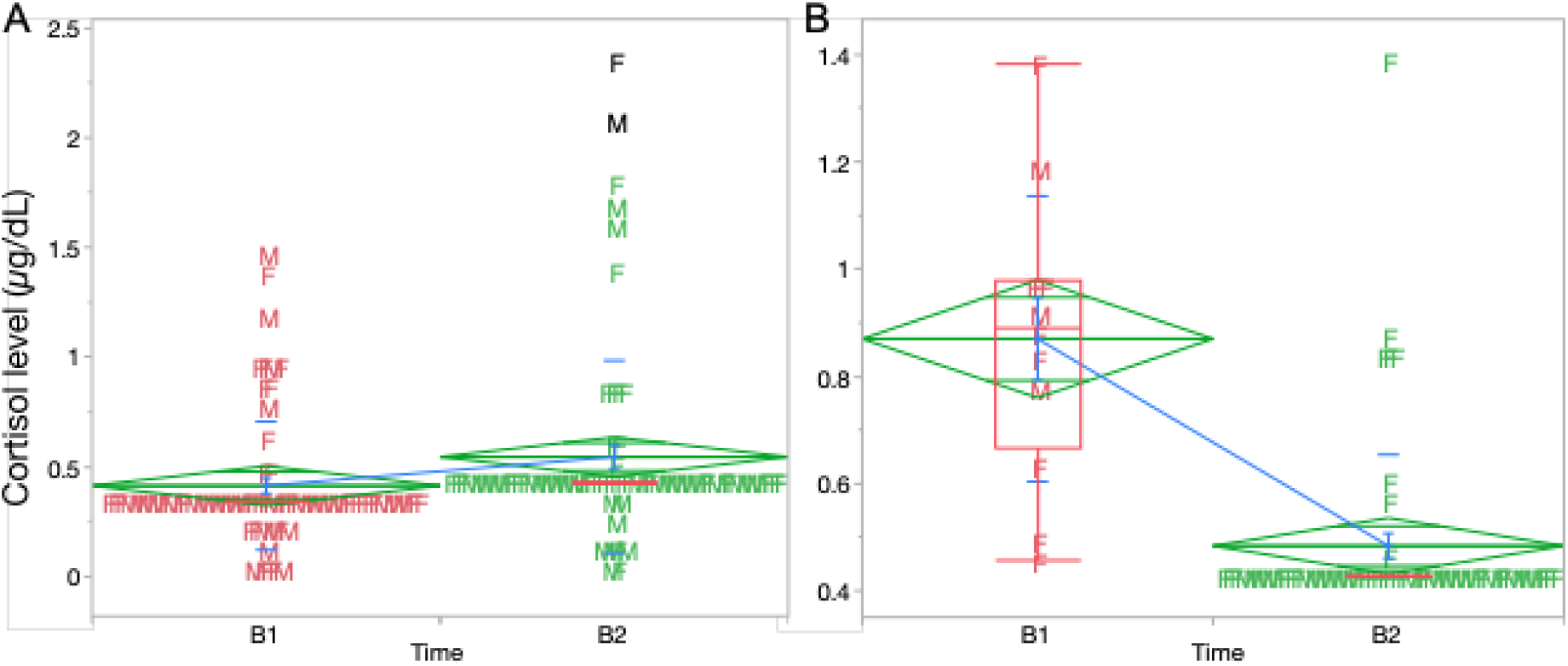
Box plots showing distribution and comparison of changes in salivary cortisol levels before (B1) and after (B2) music presentation in ICU patients. Horizontal lines indicate mean leavels and symbols indicate sex of individuals: males (M) and females (F). A. plot for all patients, both sexes; showing a slight but significant increase in cortisol levels. B. plots showing decrease in cortisol levels in the midrange (0.35 to 1.4 µg/dL) in response to music presentation in ICU patients. Red horizontal lines indicate median leavels and green diamonds represent 95% confidence intervals from the pooled stanbdard deviation. Bue line connects group means (long horizontal green lines). Short horizontal lines indicated standard deviation. Two black data points were outliers in the data.

**Figure 6:**
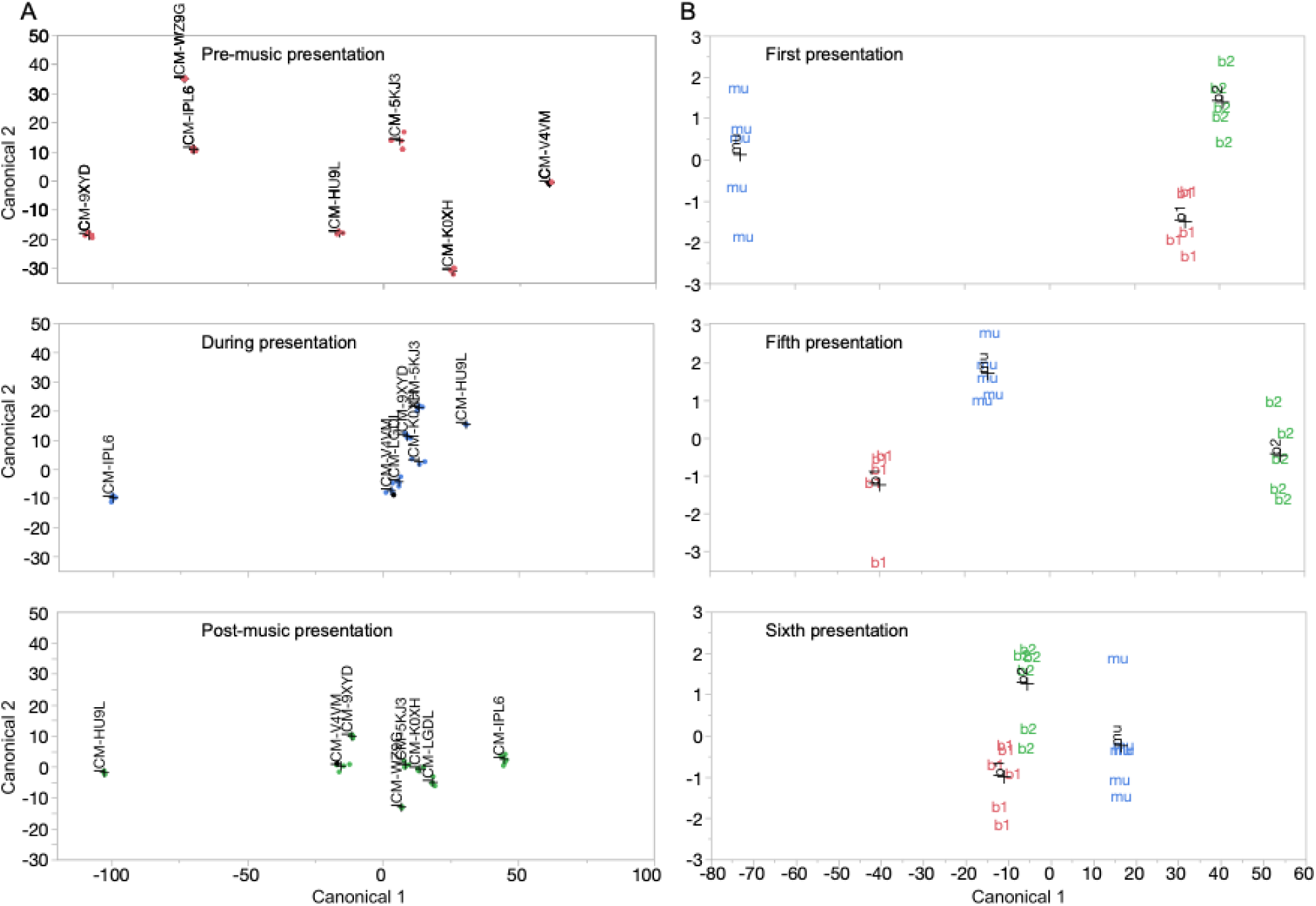
Multidimensional scaling (MDS) of twenty-five parameters related to HRV in a patient mapped all five instances of the music presentation condition outside of the mixed cluster of baseline conditions, indicating a shift (arrow) away from the baseline state.

We also performed a similar analysis to examine the cumulative effect of up to six sessions of music presentations. With session number as the discriminant, we compared the relative distance between the score for HRV parameters on the primary DCA axis for music presentation and the before and after music presentation intervals in the reduced two-dimensional space (from the multivariate HRV space). We tracked these differences during the first music presentation session with the fifth and sixth sessions in all patients (n = 8) where these data were available. The canonical details showed a good fit (P < 0.0001, F = -72.2; 110, -84 for the first DA component) and a canonical correlation > 0.99). On average the first eigenvalue captured > 99% of the variation across successive trials. The relative distance between Canonical 1 scores for the different trial-types (pre-, during and post-music presentation) decreased on average by ∼130 units. This suggested that successive music presentations induced a convergence within the HRV parameter space, driving the condition of the heart to be similar to what was induced by the first presentation of music.

We used MDS to map patient distances within two-dimensions to visualize their canonical distances from each other. We also used MDS to gauge trial-specific effects of music presentation across subjects as well as in a single subject. This patient served as a case-study for testing the cumulative effect, if any, of music presentation. These data revealed that the state of the heart during music presentation was significantly different from the baseline condition. We performed a within-subject MDS analysis to visualize the effect of music and whether there was overlap in the placement of the constellation of computed HRV parameters before during and after music presentation (Fig. 7A). The MDS plot indicates music presentation resulted in a shift in the HRV parameters away from the baseline condition. The cluster of HRV parameters during music presentation were located away from both baseline conditions, suggesting a return to the baseline physiological status of the patient post music presentation. This trend of a shift in HRV parameters during music presentation and sometimes during the post-presentation baseline condition was observed as well in other patients afforded several trials of music presentation (see Fig. S2).

**Figure 7.**
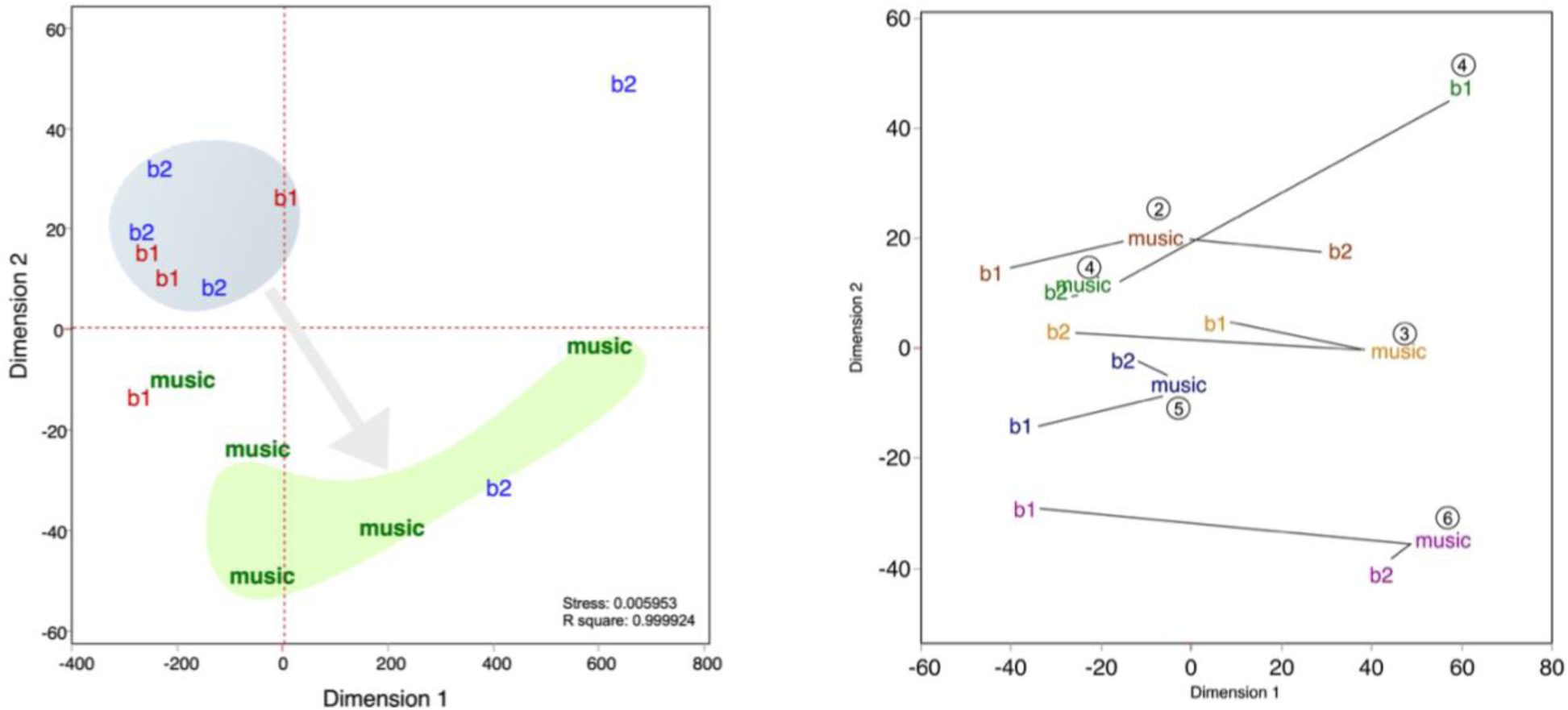
MDS of twenty-five parameters related to HRV in a single patient. A. Representation of all five instances of the music presentation condition outside of the mixed cluster of baseline conditions, indicating a shift (arrow) away from the baseline state. B. MDS plot including data plotted separately for each presentation and recording session in a subject that underwent six presentatons over a two-day period in the ICU. “b1” and “b2” are baseline conditions and “m” represents music presentation.

MDS data and plots cannot be averaged or superimposed across multiple subjects since they are obtained after rotation of the data coordinates to allow for the best representation for each subject. Therefore, we obtained a second MDS plot in the same patient in which all pre-presentation (b1), presentation (m) and post-presentation (b2) sessions were separately placed and labeled in the same MDS space (Fig. 7B). These data show a general tendency for distance between the baseline condition increased with successive music presentations to decrease with repeated music presentations, shifting the post-baseline condition closer to the music condition.

Thus, both in a single subject and across subjects, there was a shift in HRV parameters during music presentation that could have a lasting effect. This trend did not hold up for HRV parameters across subjects for when only the first and second music presentations were included in the model but persisted in results of trials where music was presented either the third or fourth time, suggesting once again that multiple presentations of music had a cumulative effect on possible improvements in the physiological status of the patient.

Finally, we performed factor analysis of the various HRV parameters to identify the parameters explaining the greatest percentage of variation in the data and to directly observe the effect of music presentation on some of these parameters. This analysis showed that the first two dimensions or factors captured ∼ 69% of the variation and seven factors captured close to 80% of the variation in the HRV parameters across all patients and all trials. A principal components analysis (PCA) identified the following parameters as the ones that most strongly define the horizontal axis in the principal components (PCs) of the PCA (i.e., spread in the HRV plot) from the HRV-based clustering:

HRV_HTI (Histogram TI)
HRV_ShanEn (Shannon Entropy)
HRV_CVI (Cardiac Vagal Index)
HRV_MadNN (Median Absolute Deviation of NN intervals)
HRV_IQRNN (Interquartile Range of NN intervals)

Some of these parameters showed a clear inflection when the patient listened to music and sometimes this effect tended to last past the post-music presentation interval recording period, i.e., for at least 15 to 30 minutes and possibly longer. (Fig. 8). In other cases, these inflections returned to the baseline post-music presentation.

**Figure 8:**
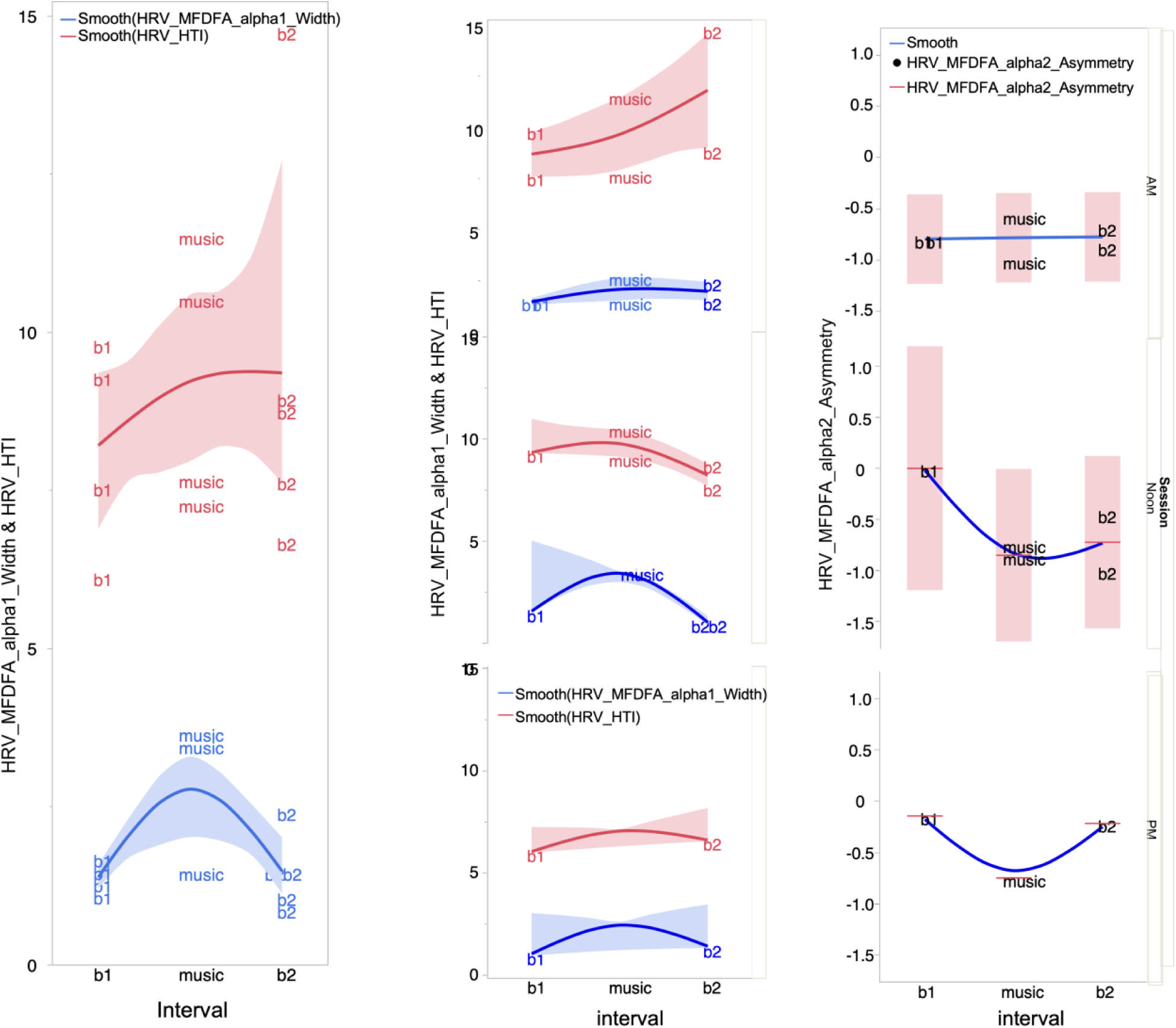
Line plots in side-to-side panles showing transient effect of music presentation at three times during the day on measures of different HRV parameters in a patient. Lines show mean values and shaded areas represent 95% confidence limits for variations.

## 5. Discussion

Our study was inspired by the possibility of obtaining data from continuous physiological monitoring with an end-goal to evaluate and personalize music interventions in the ICU as well as gain a in-depth understanding of the physiological mechanisms underlying the healing effect of music. ICU patients are typically connected to an array of monitors that record heart rate, blood pressure, respiratory rate, oxygen saturation, and sometimes advanced metrics like HRV or even EEG (in neurocritical care). These monitoring systems present an opportunity to objectively validate the real-time effects of music on a patient’s physiology. Rather than relying solely on subjective reports, we hoped to observe changes in autonomic markers during a music session. For example, a gradual reduction in heart rate and systolic pressure, coupled with increased HRV high-frequency power, during a 15-minute music intervention would strongly suggest activation of the parasympathetic relaxing response. Similarly, if continuous EEG is available, one might detect shifts in brain wave patterns – such as increased alpha or theta power indicative of relaxation – corresponding with the introduction of music. In a recent pilot study with ICU burn patients, such moment-to-moment monitoring was employed: patients showed significant EEG oscillatory changes (in delta, theta, alpha, and beta bands) during music-assisted relaxation, alongside autonomic shifts (HRV changes) and reduced facial muscle tension, even when pain and anxiety scales did not change dramatically in the short term. These findings demonstrate the feasibility of using critical care monitoring technology to capture the subtle, early effects of music on the nervous system. In short, the integration of continuous physiological monitoring with music therapy in the ICU can be a powerful approach to both understanding and validating a particular intervention for maximal benefit to the patient.

### 5.1. Clinical Effects of Music in ICU and Postoperative Care

Over the past decade, clinical research has increasingly demonstrated that presentation of music can translate into tangible benefits for patients in high-stress medical settings. The intensive care unit (ICU) environment is characterized by extreme stress, anxiety, and physiologic instability, making it an important testing ground for music-based interventions. Multiple randomized controlled trials in ICUs have documented that patients exposed to music (often via headphones with patient-selected or calming instrumental music) experience significant reductions in anxiety and distress compared to those receiving standard care (Bradt and Dileo, 2014). A review of 14 trials (805 total ICU patients) concluded that music interventions yield a large reduction in anxiety, averaging over one standard deviation unit greater anxiety relief than control conditions. In mechanically ventilated patients, the anxiolytic effect of music was accompanied by consistent physiological improvements: notably, lower respiratory rates and lower systolic blood pressure. Music presentation can also reduce sedative and analgesic requirements for ICU patients. Patients who listened to music needed significantly less sedation (and analgesia) to achieve comfort on mechanical ventilation (Bradt and Dileo, 2014). Excessive sedative use can prolong ventilation and ICU length of stay, so this is a significant benefit (Chlan et al., 2013).

Multiple studies report lower cortisol levels in participants exposed to soothing music under stress or when undergoing medical procedures (e.g., surgery). In one study, pre-stressor music listening hastened autonomic recovery (with faster normalization of salivary alpha-amylase) and tempered the cortisol response to a subsequent psychosocial stressor (Thoma et al., 2013). Such findings underscore music’s capacity to prime the body for stress attenuation via neuroendocrine pathways. By providing an alternative pathway to alleviate anxiety and agitation, music therapy may help break the feedback loop where distress necessitates sedatives, which in turn can cause delirium or other complications. It should be noted that in ICU studies measuring hormonal outcomes, results have been mixed: some have found reductions in cortisol with music, while others report no significant change, likely due to variations in timing, patient condition, and the overwhelming influence of critical illness on endocrine axes (Koelsch et al., 2011; Miluk-Kolasa et al., 1994; Uedo et al., 2004). The mixed nature of our results on effect of music on cortisol production is consistent with these observations. Effects on heart rate have also been somewhat variable across studies. Therefore, in our study we focused on heart-rate variability (HRV) rather than heart rate by itself.

### 5.2. Putative mechanisms of music-induced healing

Here we dig deeper into the neurobiological basis of music perception and healing. Music is perceived largely through the same auditory channels as all other sounds though receptors under the skin, such as Pacinian corpuscles, may also play a role in the detection of sound pressure waves. This has been suggested as a possible mechanism for sound perception of low (< 600 Hz) frequencies by those who are clinically deaf. This was a reason for us to choose a large, exceptional quality speaker that would provide a full body stimulation for low frequency sound components in music. Regardless, music eventually engages a broad neurobiological network that modulates emotion, stress physiology, and even immune function.

Pleasant music can activate the brain’s reward circuitry, particularly the mesolimbic dopaminergic system. Neuroimaging studies show increased activity in the ventral tegmental area and nucleus accumbens (ventral striatum) when individuals listen to enjoyable music. This dopaminergic surge is associated with feelings of pleasure and reward, which can counteract stress and pain reactivity. Listening to calming music has been linked to reduced activity in the amygdala that is regarded as the brain’s fear and stress center. This modulation can suppress downstream hypothalamic–pituitary–adrenal (HPA) axis activation and sympathetic nervous system output, leading to measurable reductions in stress hormones and arousal (Koelsch, 2015).

In parallel with HPA-axis effects, music exerts significant direct neural influences via the autonomic nervous system (ANS). Relaxing music typically enhances parasympathetic (vagal) activity and/or decreases sympathetic activity, manifesting as lower heart rate, reduced blood pressure, and changes in heart rate variability (HRV) indicative of a relaxation response. For instance, in mechanically ventilated ICU patients, music listening consistently reduced respiratory rate and systolic blood pressure, suggesting an activation of the vagal “rest-and-digest” response. Stress hormones like cortisol are well known to suppress certain aspects of immune function; thus, the cortisol-lowering effect of music, as confirmed by our study, may indirectly benefit immunity. Beyond this indirect link, some studies suggest direct immune changes in response to music, including a significant increase in salivary immunoglobulin A levels and significantly elevated IgA levels in addition to evoking positive emotions (Charnetski et al., 1998; Kuhn, 2002; Mccraty et al., 1996). Endogenous opioids and neuropeptides, such as beta-endorphin release has been linked to music listening. Similarly, oxytocin – a hormone related to stress reduction and immune modulation – may increase during musical engagement in group music-making settings. Feelings of social bonding can also raise oxytocin levels and dampen inflammation. The brain’s mirror neuron system may facilitate this phenomenon, though direct evidence of this is still lacking. While the precise pathways remain an area of active research, the converging data support a psychoneuroimmunological mechanism where music favorably alters hormonal and autonomic outputs, reducing perceived stress and negative affect. This in turn can lead to measurable changes in immune markers (e.g., immunoglobulins, cytokines, or natural killer cell activity).

Finally, music’s ability to capture attention and emotion can also serve as a cognitive distraction, diverting patients’ focus away from pain or worrisome thoughts, thereby breaking the vicious cycle of anxiety and physiological arousal. Through this combination of reward activation, limbic calming, and autonomic adjustment, music creates a neurochemical milieu that may promote healing. Engaging the mirror system might enhance social connectedness. Patients who participate in music therapy often report feeling understood or supported, possibly because the music creates a sense of interpersonal synchrony with the therapist or composer. As a multisensory, embodied experience, music listening does not involve merely processing of sounds by the auditory cortex in isolation, but an experience that involves activation of memory and emotional (limbic) networks and movement networks. Thus, music listening engages a network of brain regions (reward, limbic, cognitive) that jointly regulate the HPA axis and ANS, thereby translating emotional experiences into peripheral physiological effects on stress and immunity.

### 5.3. A “Healing Network” in the embodied brain

The limbic-autonomic integration discussed above provides a framework for how music’s effects propagate from the brain to the body. Activation of cortical and subcortical emotional centers (e.g. the orbitofrontal cortex and amygdala) can modulate brainstem autonomic nuclei (like the vagal nerve complex), leading to changes in heart, lungs, and adrenal glands. Simultaneously, reward-related neurotransmitters (dopamine, endorphins) and neurohormones (oxytocin) triggered by music can enter the blood circulation and signal immune organs (spleen, bone marrow) to boost immune cell activity. This suggests that music might engage a neuroimmune network, resulting in downstream signals that reduce pro-inflammatory cytokine production and enhance immune regulation. Indeed, some studies have noted changes in cytokine levels (e.g., interleukins) associated with music or singing interventions in clinical populations (Fancourt et al., 2016). Though more research is needed to map these connections explicitly, current data align with the concept that music orchestrates an ensemble of brain regions and bodily systems (see Fig. 8). By simultaneously influencing the emotional brain, the autonomic nerves, the endocrine glands, and the immune defenses, music listening bridges one’s psyche and soma. Incorporating or taking advantage of such a network can have profound implications for holistic patient care.

Building on the above concepts, we propose the existence of a **“**healing network**”** – a coordinated system of neural and physiological elements that are jointly activated during music therapy to promote recovery. This concept serves as a theoretical framework to understand how music’s diverse effects converge to facilitate healing. The healing network is not a single anatomical pathway, but rather a functional integration of multiple pathways within the brain and those looping through the body. It encompasses the limbic reward circuit (providing positive affect and motivation), the stress-regulatory circuit (including amygdalar inhibition and HPA-axis modulation), the autonomic circuit (balancing sympathetic and parasympathetic outputs), and the socio-cognitive circuit, including mirror neuron and default mode networks that foster social connection and meaning. When a patient listens to therapeutic music, these circuits engage in synchrony – like different instruments in an orchestra playing a harmonious piece – resulting in a state conducive to healing: the patient feels emotionally comforted, physiologically relaxed, and perhaps spiritually uplifted or connected.

### 5.4. Practical implications and future directions

Our results reported here together with work of others suggest that music exposure can function as a viable, low-cost adjunct to standard medical care in settings like ICUs and surgical recovery units. Hospitals can implement programs to provide music to patients during particularly stressful times – for example, immediately after major surgery, during mechanical ventilation or bedside procedures, and even during preoperative waiting periods. Implementation can range from simple (providing patients with headphones and access to a library of pre-selected calming music or facilitating use of their own preferred music) to more structured scheduled sessions with a certified music therapist who can tailor the intervention to the patient’s as well as a long-term care provider’s needs. Given the heterogeneity in patient music preferences, allowing patient-selected music is generally advisable, as familiar and favorite music may amplify the positive emotional response. However, in cases of severe anxiety or agitation, professionally curated music, as used in this study, with a combination of arousing and soothing melodic structure, depending on the time of the day, might be optimal to promote relaxation and healing. Training ICU staff about the potential benefits of music and offering but not forcing interventions will be respectful of individual differences.

## 6. Conclusions

In conclusion, the role of music in medicine, particularly in modulating the brain, behavior, and immunity of patients under high stress, is supported by a growing body of scientific evidence. Through neurobiological pathways involving the limbic system, autonomic regulation, and possibly direct neuroimmune interactions, music can induce a state of relaxation and positive emotion that translates into better clinical outcomes. As research continues to unravel the “healing network” activated by music, we move closer to a future where caregiving involves not just pharmaceuticals and procedures, but also the profound, empathetic power of art and sound to heal the human body and spirit. Finally, continued innovation in monitoring and personalization will enhance the efficacy of music therapy. Continuous monitoring, as was originally a goal of our study, not only can provide evidence of effect but can also guide intervention adjustments. If a patient’s heart rate does not decrease with a given musical piece, clinicians might try a different genre or tempo to find one that resonates more effectively with that individual’s physiology. This real-time feedback loop could herald a form of personalized music therapy, where biosignals inform the selection or modulation of the auditory stimulus, analogous to how anesthesia dosing is adjusted based on vital signs. Moreover, continuous data collection enables robust analysis of dose-response relationships – for instance, determining how long a music session should last to achieve a sustained reduction in stress indices, or how quickly physiological benefits emerge after music begins.

## 7. Ethical standards

Our study met all ethical standards typically prescribed for clinical studies.

## 9. Credit authorship contribution statement

Conceived, designed and supervised the study J.S.K. and J.L.), Music creation (A.S.), Methodology development and implementation (J.S.K., J.L., J.M. P.K. A.J.P. and A.P.), data analysis (J.S.K. J.M. A.J.P. and A.P.), J.S.K. wrote the original draft of the manuscript. All authors reviewed, contributed to and approved the final version of the manuscript.

## 11. Data availability

Data can be made available upon request post publication.

## 12. Acknowledgements

We thank Drs. Rohit Satoskar, MD, and Eleanor Drew, MD for their enthusiasm and willingness to allow their patients to have the option to be exposed to music listening sessions post-surgery and Sunil Iyengar for his interest and openness to consider our study for funding by the NEA panel of scientists and artists. We also thank all the patients who chose to participate in this study and make this work to be possible.

